# Co-production of informal settlement health: A community based participatory research program for building healthy communities in urban informal settlements of Salvador, Brazil

**DOI:** 10.1101/2025.02.19.25322540

**Authors:** Hammed Mogaji, Lopez Yeimi Alexandra Alzate, Lívia Almeida Figuerêdo, João Henrique Araujo Virgens, Marie Agnes Aliaga, Hernan D Argibay, Inajara Salles, Andreane Pereira Moreira, Terezinha de Jesus Lima e Silva, Suzana Cristina dos Santos, Rita Batista, Elizete Cardoso, Elenilda Cardoso Neves Santos, Edlane Leal dos Santos, Edlana Rodrigues dos Santos, Thiago da Mata Barreto, Thais Auxiliadora dos Santos Mattos, Nivison Nery Júnior, Jaqueline Cruz, Ianei Carneiro, Ricardo Lustosa, Victoria C. Dedavid Ferreira, Mitermayer Reis, Albert I Ko, Federico Costa, Mike Begon, Hussein Khalil

**Affiliations:** Instituto Gonçalo Moniz, Fundação Oswaldo Cruz, Ministério da Saúde, Salvador, 40296-710, Brazil; Instituto de Saúde Coletiva, Universidade Federal da Bahia, UFBA, Salvador, PO BOX 40110-040, Brazil; Department of Epidemiology of Microbial Diseases, Yale School of Public Health, New Haven, USA, 06511; Association of Residents of Alto do Cabrito Salvador, 40484-210, Brazil; Cia de Arte Cultural E ao Quadrado, Salvador, 40484-550, Brazil; Associação Emília Machado Bahia, Salvador, 41280-000, Brazil; Department of Preventive Veterinary Medicine and Animal Production, Escola de Medicina Veterinária e Zootecnia, Universidade Federal da Bahia, UFBA, Salvador, Brazil; Faculdade de Medicina da Bahia, Universidade Federal da Bahia, Salvador, Brazil; Department of Evolution, Ecology and Behaviour, University of Liverpool, Liverpool, L69 7BX, United Kingdom; Swedish University of Agricultural Sciences, Umeå, Sweden

**Keywords:** Brazil, Health inequities, Urban slums, Health justice, community-based participatory action research, Popular Health Education

## Abstract

More than 15% of Brazil’s urban population lives in slums characterized by limited access to essential urban services, heightened vulnerability to infectious pathogens and environmental hazards, and deprivation of citizenship rights. These conditions exacerbate social inequality, perpetuate cycles of poverty, and fuel violence, underscoring the urgent need for sustainable interventions. Following a social justice framework, we developed a community development program rooted in participatory research methods and popular health education to foster collaboration between university researchers and communities. The aim was to identify priorities and co-create locally driven, cost-effective, and sustainable solutions. This article describes our ongoing project in three prominent urban slums of Salvador, Brazil (Alto do Cabrito, Pau da Lima and Marechal Rondon), detailing the methodologies employed, activities initiated, and interventions developed. We conducted ethnographic, eco-epidemiological, and collaborative mapping surveys to contextualize diverse health and well-being challenges. Furthermore, we organized consultative and socialization events with dynamic community groups identified local priorities, leading to the design of 13 interventions targeting citizenship rights, social cohesion, environmental restoration, waste management, and unemployment. Here, we described how our interdisciplinary approach leveraged social capital and fostered inter-sectoral partnerships to empower marginalized urban communities towards addressing their health and environmental challenges through sustainable, locally tailored solutions. While the program has strengthened community trust, facilitated partnerships, and achieved notable environmental improvements, further evaluation is needed to assess the long-term impacts of these interventions on broader social health determinants.

## Introduction

Over a billion people reside in informal settlements within urban areas, with a projection to rise by more than 300% by 2030 [1]. These settlements are characterized by inadequate housing conditions which impacts urban equity and inclusion, health and safety, and livelihood opportunities [2]. Brazil, like many other low-middle income countries, faces the challenge of high levels of social and economic inequalities in urban areas [3,4], resulting in the establishment and expansion of urban informal settlements, commonly referred to in Brazil as “*favelas*” [5]. More than 15% of the Brazilian urban population resides in *favelas*, where they have limited economic opportunities, insufficient access to basic urban services, including clean water and sanitation, and health care [6]. Informal settlement residents often live in substandard and overcrowded domiciles, subject to seasonal flooding and erosion risks, which increases the exposure to pathogens whose transmission is favoured by such poor socio-economic and environmental conditions [7,8,9,10]. Social inequality, health disparities, and in some cases, violence in informal settlements are closely linked to feelings of deprivation of citizenship rights and privileges [11].

Over the last two decades, our team has been engaged in the study of infectious disease in residents of urban informal settlements in the Brazilian city of Salvador, Bahia [7–10]. Initially the focus, led by medical scientists, was on the epidemiology of leptospirosis, a neglected but important human bacterial disease transmitted from the urine of a zoonotic reservoir host, predominantly the brown rat, *Rattus norvegicus* [9,10,12,13,14]. Subsequently, our scope broadened to encompass ecological studies of leptospirosis in its *R. norvegicus* reservoir [15], and in the wider environment [16], and to other infectious diseases, including dengue [17], zika [8], and SARS-CoV2 [18]. Recently, the research program broadened further to fully incorporate inputs from social scientists, to promote community engagement and participation [19, 20], and to include health and wellbeing more generally and their social, demographic, and environmental determinants [21].

Collectively, our studies have highlighted the need for investments in environmental improvements, basic urban services, and enabling residents to protect themselves and their domiciles from sources of environmental contamination [11]. For example, investment in infrastructural changes and improved trash collection as a potential strategy to reduce reservoir and vector populations, environmental contamination, and contact between residents and pathogens exposure to pathogens has continued to gain support [10]. However, such transformations demand significant government investments, which, in practical terms, may not materialise within an expected timeframe. Instances of political neglect and disparities in the allocation of infrastructural resources to informal settlement populations are not uncommon [22], leading often to sporadic and localised interventions that lack coordination, and which are therefore unsustainable. Sustainability is further undermined by interventions often being externally imposed, short-term, and lacking contextual relevance and so being prone to issues of acceptability, appropriateness, and feasibility [23].

These considerations, and the growing complexities associated with urbanisation and inequality globally, argue for the need, far more widely acknowledged in the social than in the natural sciences, for teams such as ours to adopt approaches that fully involve community residents in the creation and delivery of interventions tailored to the specific context of informal settlement areas. This necessitates a recognition of professional researchers’ social obligation and commitment to research processes that can drive sustainable change [24], predicated on the premise that co-construction of knowledge between professionals and communities allows for more synergistic relationships and deeper engagement, which may be key not only to health promotion, but also to social transformation and emancipation in these informal settlements [25,26].

Researchers are increasingly turning to available participatory and implementation-driven research perspectives to address health disparities stemming from socioeconomic and environmental disadvantages [27,28]. Our on-going program, described in this article, therefore adapts and builds on the principles of Community-Based Participatory Research (CPBR) and Popular Health Education, which entails conducting research in collaboration with the communities affected by the object of investigation, with the primary aim of driving action and facilitating social change based on local priorities [29,30]. We identified several key attributes for the success of this approach, including successful blurring of lines between researchers as neutral and objective observers and research participants, minimizing power imbalances, approaching health and wellbeing with a holistic and community-based perspectives, and conducting research in partnership with communities to achieve common goals: sustainable, long-lasting positive outcomes that extend beyond the duration of the research [29]. Our aim is to address the paucity of CBPR approaches necessary for improving health outcomes among informal settlement populations [22,31]. Hence, we worked with residents and other stakeholders in three communities that are superficially similar, but have contrasting histories and social characteristics, leading to three unique CBPR experiences. These included a range of participation dynamics and community empowerment activities and outcomes, leading to cross-fertilisation arising from the interaction among communities and our interdisciplinary research team. Nevertheless, a common aim in the three communities was to address simultaneously multiple problems of health and well-being (a range of infectious diseases, rat infestation, food security, and environmental and community wellbeing) and evaluate multiple outcomes related to the participatory and organizational processes, stakeholder satisfaction, and individual, community, animal, and environmental health.

Hence, in this article, we describe the processes and results achieved in the initial phase of our program. In doing so, we draw an important distinction between what we describe as “activities” and “interventions”. Indeed, the feedback between activities and interventions is a key characteristic of the approach described here. We define an activity as an action that is initiated by the external research team (i.e., *not* by the community), which has the proximate aim of facilitating community engagement or empowerment and can thus be assessed by its success in doing so, and has the further aim of generating, through interactions between the external research team and the community, one or more interventions. On the other hand, an “intervention” is defined as an action initiated by an empowered and engaged community in collaboration with the external research team, with the aim of improving the health and/or the wellbeing of the community and can thus be assessed by its success in doing so in the short and/or the long term.

In terms of presentation outline, we first provide the health context of the study area and described the range of *activities* undertaken in the interdisciplinary and CBPR process with the aim of developing sustainable community-owned interventions towards wellbeing and empowerment. This is ‘what was done’ and might therefore be considered equivalent to our ‘Methods’. We then describe the proposed *interventions* generated by those activities. The interventions are the outcomes of what was done, which might therefore be considered our ‘Results’. Finally, we discuss several key findings and lessons emerging from reflections on the CBPR process leading from activities to interventions, including how they differ from the conventional solutions health experts might have suggested, and how their success is to be assessed. Such assessment will be described in subsequent articles.

## Methods

### Study area, population and context

Our program was implemented in the city of Salvador, state of Bahia, North-eastern Brazil, where more than 2.6milion people resides [6]. In Salvador, 45% of households live in informal settlements [6], and the city has about 7% of all urban informal settlement residents in the country [32]. Most of our previous studies have been implemented in the Pau da Lima community [8,9,18], and more recently in four other similar marginalized peri-urban settlements including two considered here, namely Marechal Rondon and Alto Do Cabrito [10,19]. All these communities are extremely vulnerable to flooding and soil erosion due to their open sewer systems and their topographic properties, including steep valleys (Figure 1). In these communities, most residents lack legal titles to their homes, and the median household per capita income is US$2.32/day [18]. During the COVID-19 pandemic, economic conditions pushed more than two-thirds of the population into hunger and close to 80% into unemployment [10,33]. The communities differ in age of establishment, yet variation in socioeconomic and environmental conditions appears to be larger within than between the communities [10,33]. Within each community, our study area ranges between 0.07 and 0.09 km^2^ in extent, and the project has approximately 3000 participating residents, living in over 1000 households (see Table 1 for the information on other data collected).

**Figure 1:**
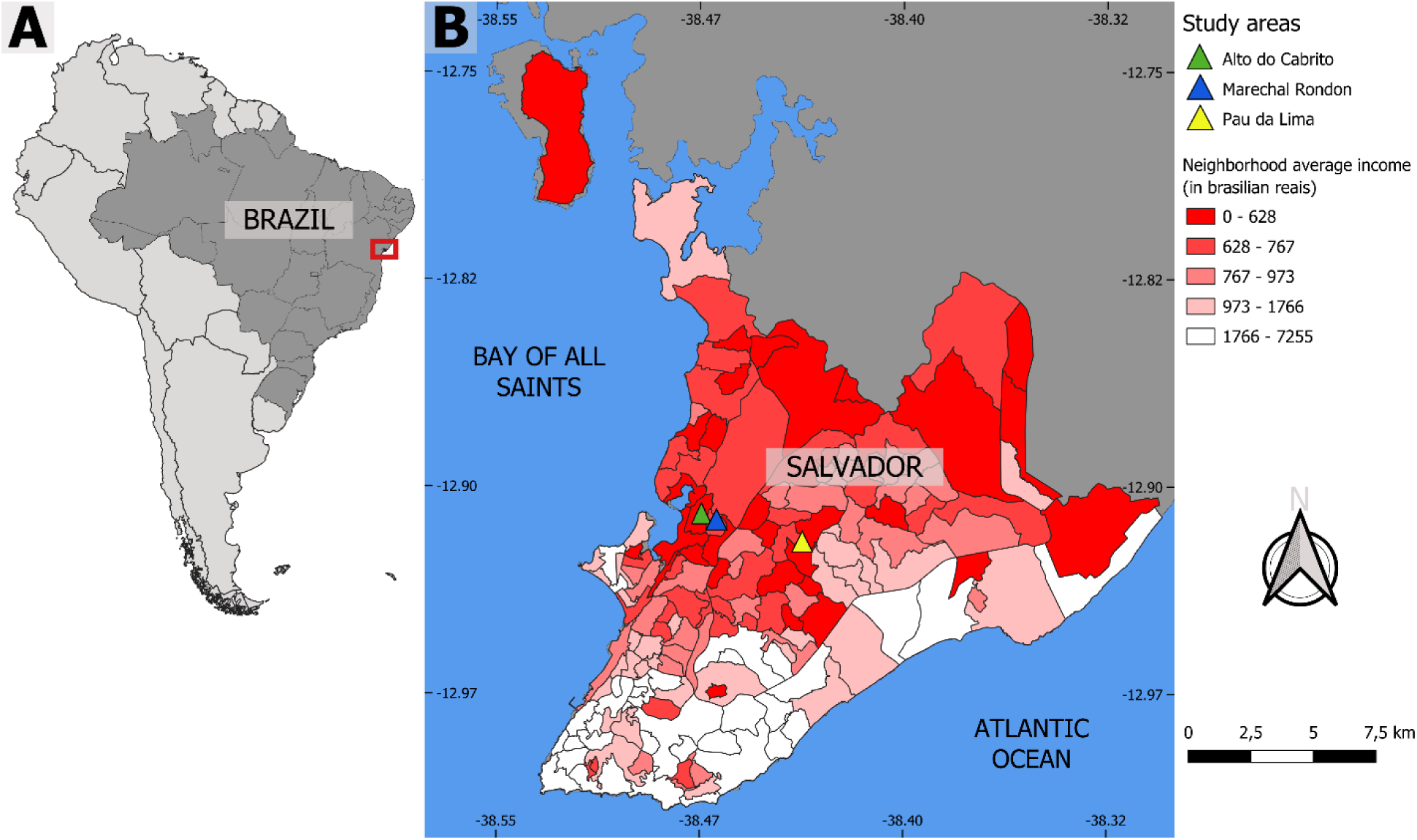
Map of Brazil and Salvador showing the location of the three marginalized communities where we work. The neighbourhoods of Salvador are classified by the mean income of the head of the household to show the rent spatial segregation observed in the city. The maps are not copyrighted as they were created by the authors using QGIS 2.18 software. The boundary polygons of Brazil, Salvador and the production of the Salvador income map were downloaded from open and publicly accessible base of IBGE—Instituto Brasileiro de Geografia e Estatistica, on the Geosciences platform which can be accessed at https://www.ibge.gov.br/geociencias/downloads-geociencias.html. The WorldView-3 May 2017 satellite image was also used to digitize the study area. The image was acquired by the research project / Instituto Gonçalo de Moniz—IGM—Fiocruz Bahia from the company Satmap, with disclosure permitted referencing the Copyrights of DigitalGlobe images.

**Table 1:**
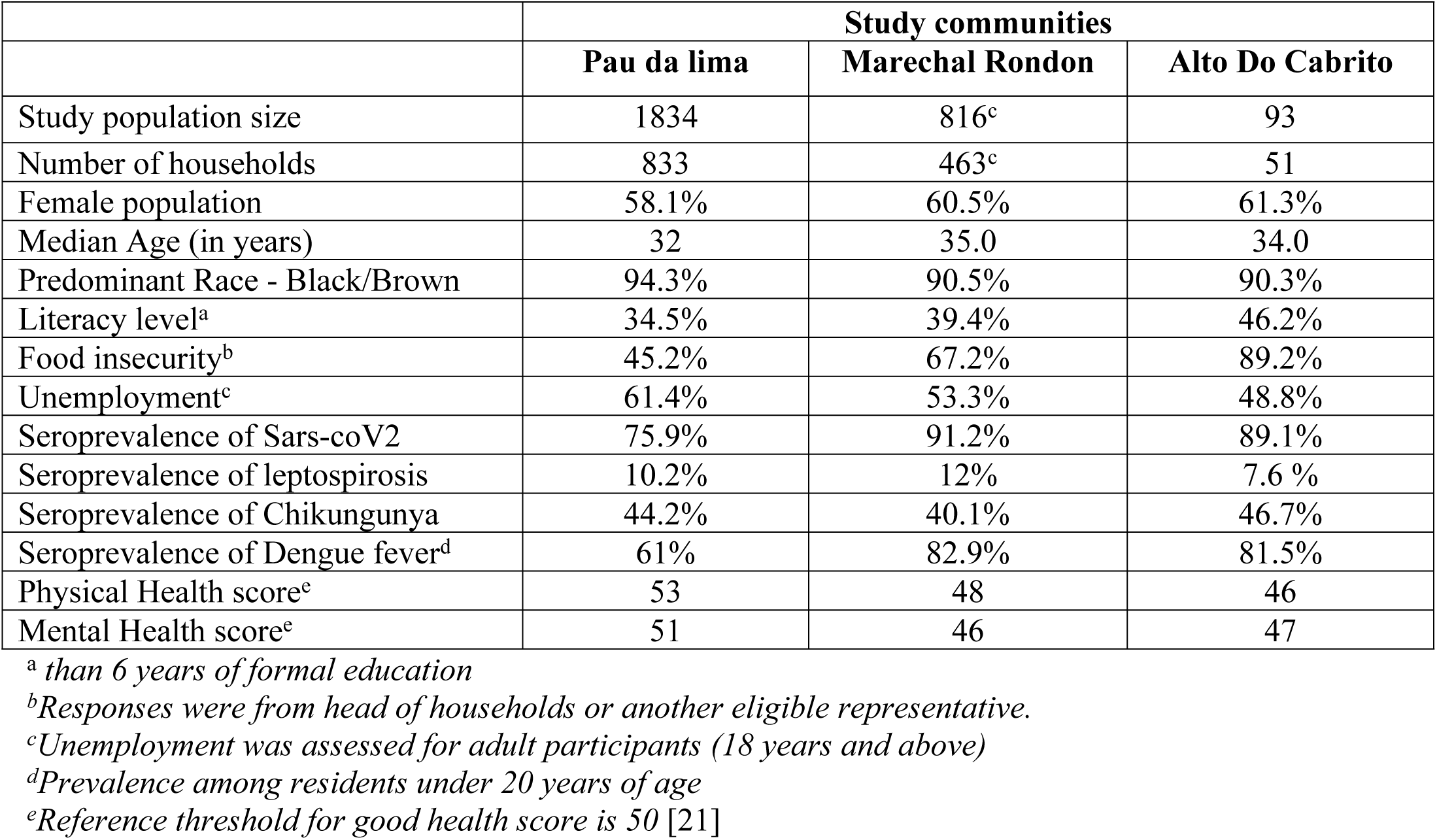
Baseline profile of study communities included in the interdisciplinary project (November 2021)

### Program design

To integrate principles of popular health education into CBPR, our design includes the following dimensions: (i) community engagement (events and activities) to facilitate the entry and participation of community residents and other stakeholders, (ii) quantitative, eco-epidemiological research to estimate exposures of residents and domestic animals to disease reservoirs and vectors, as well as collaborative environmental and perception mapping to provide accurate geo-location of the problems identified by the communities. Additionally, the project includes (iii) qualitative research and ethnographic studies to understand the processes of participation and the perceptions of residents about the various processes of research, (iv) socialization activities for co-construction of idea and decision making, (v) dialogue and cooperation with local agencies (public and private) for the implementation and possibility of scaling up the proposed interventions, and finally (vi) co-evaluation of research processes with communities, which includes quantitative and qualitative parameters. (Figure 2)

**Figure 2:**
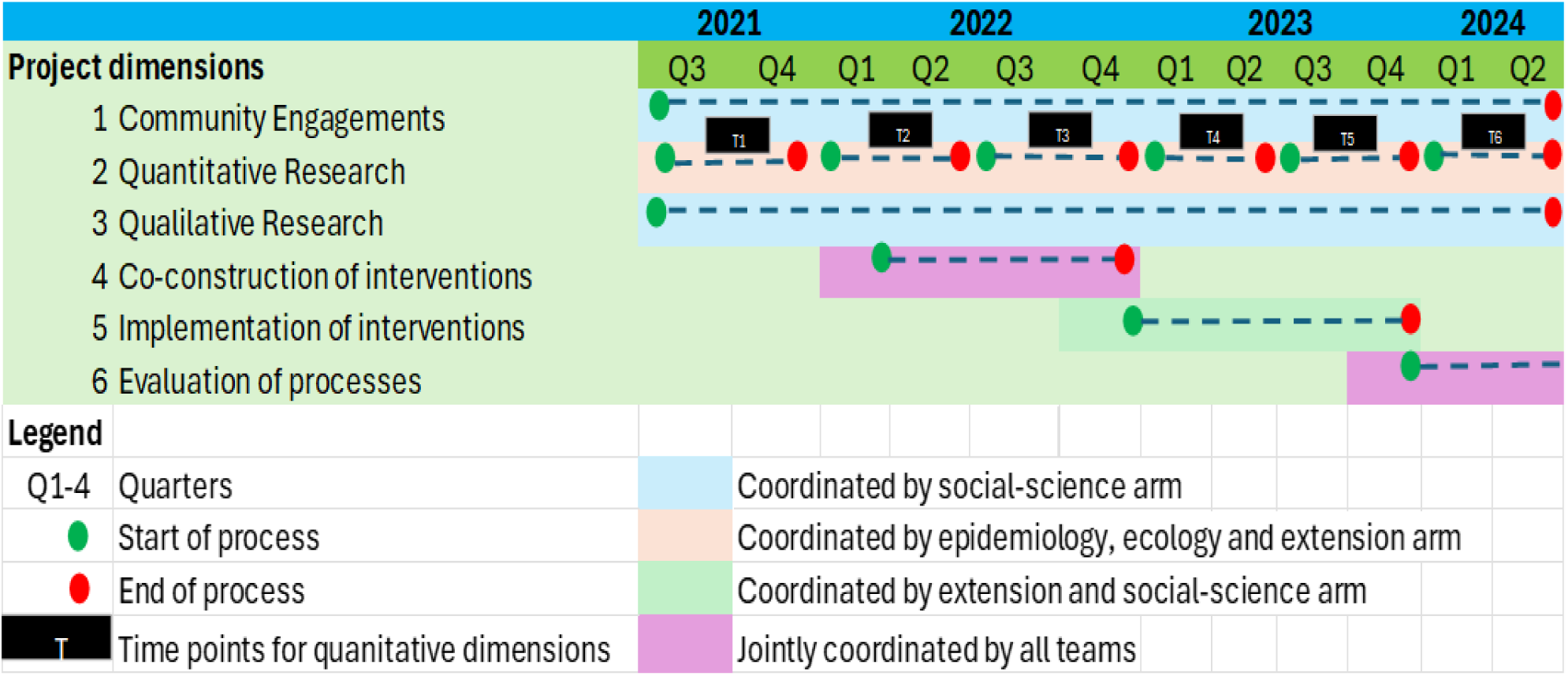
Schematic diagram of the project design

### Program timeline

The program has a three-year timeline, and of the methodologically diverse dimensions listed above, (i) community engagement, (iii) qualitative research, and (iv) engagement and mobilization started at the inception of the project and continued throughout its lifetime. In contrast, for the eco-epidemiological, intervention implementation, and process evaluation dimensions ((ii), (v), and (vi)), there were six approximately bi-annual timepoints (T1 – T6). For the first six months (T1 until T2), all arms of the project-initiated baseline activities. Between timepoints T2 and T3, community engagement and the socialization of previous and baseline (T1) results were carried out to facilitate discussions regarding intervention development, marking a pre-intervention phase extending from T1 to T3. Subsequently, interventions were initiated at T4, and an intervention phase will extend between T4 and T5, while the evaluation of interventions will extend up to T6.

### Team composition and role

The program has three interlinked arms: (i) social sciences, (ii) ecology and epidemiology (eco-epidemiology), and (iii) collaborative mapping and extension. Briefly, the social sciences arm is coordinated by a team of social scientists, educators and psychologists. They organize socialization activities including engagement within and between teams and residents, perform ethnographic surveys, and subsequent curation and dissemination of research findings. All team members were trained at the outset in the concept of popular health education prior to community dialogues. The training combined theoretical and practical experiences focusing on oppression, emancipation, and community development through grassroots work.

The eco-epidemiological arm is coordinated by a team of epidemiologists, ecologists, statisticians, database managers and laboratory technicians. They perform and supervise surveys between 01 November 2021 to November 2024 to collect demographic, socioeconomic, and environmental data as well as estimate resident and domestic animal exposure to infectious diseases. Written consent was obtained from each participant at the household level. For minors, assent was obtained using thumbprints, alongside written consent from a legal representative, usually their parents. The collaborative mapping and extension arm oversees mapping activities, focusing on resident perceptions of, and attitudes regarding, their direct (peri)domestic environment and community, including areas and environmental features they prioritize for interventions. This complements the qualitative and survey-based activities of the two other project arms. Additionally, this arm leads capacity-development initiatives within the community through institutional collaborations and engagement with governmental and other third-party organizations.

While each project arm took the lead on one or more of the project dimensions (social sciences led dimensions (i) and (iii), the eco-epidemiology team led (ii), and the collaborative mapping and extension team led (v)), the participatory dimension (iv) and the evaluation of all interventions and the process (vi) were jointly led. More importantly, even for “disciplinary” dimensions, continuous dialogue among different teams and the exchange of experiences, ideas, and mutual curiosity ensured that our project constantly evolved and interdisciplinarity increased. This is exemplified by the co-construction of novel study tools, generation of ideas, and subsequent implementation of collaborative activities among the arms.

### Recruitment and training activities across project arms

Qualified technicians and research assistants were recruited and trained using protocols and standard operating procedure tools developed specifically for each arm of the project, but also for the overall aims of the project. Further, the social science team laid the foundation for training all project members and community stakeholders in the concepts of participation and popular education, focusing on processes of oppression and possibilities for liberation, prior to dialogues carried out with the community [34,35,36]. The training combined theoretical instruction with field experience. All arms had regular bi-annual retraining activities, and weekly meetings to reflect on challenges and successes through the lifetime of the project.

### Formation of community working groups by the social science team

The primary strategy for engaging residents in the three study areas, especially for designing and implementing interventions, was the establishment of community groups. The social sciences team developed community mobilization processes (detailed below for each community), facilitated the formation of working groups, and participated in all meetings and decision-making in an equitable way with the communities. The meeting dynamics followed popular education methodologies and a problem-solving approach, allowing residents to decide on discussion topics and actions to be undertaken in their communities. Residents were regarded as active contributors to the co-construction of knowledge and social action. Table 2 provides an overview of three study communities, highlighting key characteristics of existing community groups, the target population, group size, methods employed in contextualizing needs and priorities, methods of stakeholder invitation, choice of meeting venues, meeting frequency, format, and outcomes of group discussions.

**Table 2:** Profile of community groups across the study communities included in the interdisciplinary project (November 2021)

|  | **Study communities** |  |  |
| --- | --- | --- | --- |
|  | **Pau da lima** | **Marechal Rondon** | **Alto Do Cabrito** |
| Were there existing community groups? | Yes<sup>a</sup> | Yes<sup>b</sup> | Yes<sup>c</sup> |
| Make-up of group | Mostly women and children and researchers | Community leaders, youths and researchers | Women (community leaders, residents, youths) and researchers |
| Size of group | Dynamic<sup>d</sup> | Dynamic<sup>d</sup> | Dynamic<sup>d</sup> |
| How we contextualize needs | Open questions in the sero-survey and discussions held in the Freirean tent | Open questions in the sero-survey and conversation circles with young people and leaders based on data from previous research and other problematization | Open questions in the sero-survey and conversation circles with community leaders, youths and leverages on previous research in the locality |
| How group members were invited | Door-to-door visits to residents. Delivery of printed invitations, and wall posters in the community | Contact with AEMBA and recruitment at the local school | Contact with E<sup>2</sup> and a snow-ball sampling approach among community leaders/stakeholders |
| Meeting venue | A newly created open tent by the team called a “Freirean Tent” | AEMBA and Community School | E<sup>2</sup> Theater Cultural Center |
| Frequency of meetings | Fortnightly | Weekly | Bi-weekly |
| Format of meetings | Physical | Physical and virtual | Physical and Virtual |
| Themes of group discussions | Community needs, problems and aspirations and definition of interventions | Community needs, problems and aspirations and definition of interventions | Community needs, problems and decision on priorities in interventions |

*a: Yes, but in the serosurvey questionnaire, people indicated that they did not feel represented by* *them* *b: Emilia Machado Association (AEMBA)* *c: E<sup>2</sup> Theater Cultural Center* *d: Participation is open, hence the groups are heterogeneous and dynamic in nature.*

For instance, the Pau da Lima community is very heterogeneous, characterized by its diverse composition of four valleys. The decision to work in the first valley “Baixa de Santa Rita” stemmed from three primary considerations. Firstly, the area exhibited heightened social vulnerability, notably as it initially lacked basic sanitation interventions. Secondly, epidemiological studies highlighted a significant concentration of leptospirosis cases in the region [8,9,18]. Lastly, unpublished feedback gleaned from sero-survey questionnaires revealed a sentiment among residents that they were not adequately represented by neighbourhood associations and leaders. Additionally, there was a notable absence of leisure activities or avenues for community participation reported by individuals in the area. Hence the team conducted door-to-door visits and discussions to contextualize needs, resulting in a greater participation of women who arrived with their young children to join group discussions. The team established a common ground between two areas of the community that had not seemed previously to communicate (due to geographical and social reasons), where an open tent called the “Freirean Tent” was situated, serving as the meeting venue every fortnight. To foster engagement and inclusive dialogue, we employed methodologies rooted in popular education and participatory research, including social cartography [37]. This approach enabled the identification of community needs, challenges, and aspirations for a better future. Interestingly, the involvement of children under 12 years old was unplanned initially, but mothers arrived with their children seeking participation in the sessions. Consequently, we adapted by organizing parallel recreational, sports, and artistic activities for the children, alongside the main discussions. These gatherings facilitated dynamic conversations on potential intervention strategies that could be prioritized to address community issues (Table 2).

In selecting the participatory group for the Alto do Cabrito community, we drew upon our previous collaboration with the “Cia de Arte Cultural E ao Quadrado” (E^2^), established during research conducted from 2017 to 2019 [10]. This pre-existing relationship fostered a sense of trust between the E^2^ leaders and our project team, facilitating continued collaboration. (Table 2). In Marechal Rondon, like Alto do Cabrito, there existed a prominent Community Association known as “Associação Emília Machado Bahia” (AEMBA), which we had engaged with in previous projects. AEMBA focuses primarily on youth capacity-building initiatives. To understand the community’s needs, we conducted key informant interviews with local leaders and drew upon knowledge gained from our previous research in the area. Youth recruitment for group discussions took place at AEMBA, and meetings became more frequent with the initiation of the current project. The discussions led to collaborative mapping exercises and the identification of key themes by young participants from AEMBA and Colegio Germano Machado, a school in the community, which subsequently informed action plans (Table 2).

### Activities of eco-epidemiological, and collaborative mapping arms of the project

The eco-epidemiological arm conducted surveys to estimate zoonotic and mosquito-borne pathogen burdens and associated risk factors in the study areas. For individuals above the age of 5, bi-annual serological surveys were conducted at households across study communities, involving the collection of blood, nasal swabs, and saliva (from humans). For domestic animals (e.g. dogs, cats, chickens), blood, oral/rectal swabs and feces were collected to assess health conditions and zoonotic infections. Ecological assessment involving seasonal environmental surveys using track-plates and oviposition traps were used to estimate rodent and mosquito populations [38,39]. Remote sensing techniques were also used to assess the presence and extent of open sewers, drainage, trash accumulation points, and land cover/land use. Water from open sewers and puddles, as well as soil from randomly selected points, were collected to quantify pathogen loads in the environment. These data were complimented by those provided by the mapping and extension team, which coordinated all project-related mapping activities, focusing on precise geo-location of households and environmental features that were identified by residents as points of strength or which needed improvement. The team also emphasized community empowerment through the establishment of various cultural, artistic, and communication programs. They also actively engaged with government agencies and external organizations to provide support for proposed infrastructural development projects.

### Socialization of results and development of interventions

All socialization activities (communicating and reflecting on the current state of data-based knowledge to the broader community) happened after the first 12 months of initiating the project, which coincided with the period where results generated from established working groups, and eco-epidemiological surveys had been analysed for dissemination at community meetings. The process of socialization was different in each community. The Alto Do Cabrito community served as the pilot site for socialization of results, which involved several workshops and engagements between the team and the residents over a period of 4 months. The workshops provided a platform for our team to share their research findings, enabling interdisciplinary discussions among experts in epidemiology, ecology, extension, and social sciences. The workshop methodology involved community debates based on a video synthesis of research results, mediated by field researchers with active participation from those who worked on the database, even if they had not previously visited the community. The workshop spanned two consecutive weekends, focusing on community feedback regarding what they learned from the results and how they perceived the implications for health interventions in the community. Discussions related to research results also addressed divergences between individual and structural perspectives in addressing health-care issues in the neighbourhood.

In Marechal Rondon, the socialization was not limited to meetings, but also a series of engagements between researchers and youths through training activities, field tours and mapping of identified vulnerabilities. The pre-existing interest of the youths in activities related to environmental health, vector and reservoir ecology allowed the exchange of knowledge and research findings to be more fluid and constant throughout the project.

In Pau da Lima, the socialization process took on a distinct dynamic driven by the longer-term engagement of the broader research team with the community. The project teams recognized the importance of not only sharing the results from the initiation of this project with the community, but also proposing to review published articles from research studies conducted in Pau da Lima over the last two decades. All teams, including community mobilizers, participated in this process, with the aim of organizing key points from these studies and producing written or online materials. The goal was to create preliminary versions for community dialogue and assessment, ensuring alignment with the community’s interests. However, as we were finalizing the article summaries, a sewerage improvement project commenced, disrupting our regular fortnightly meetings in the Freirean tents. During our meetings, we emphasized the connections between our research findings and the sewage improvements, and this enriched the dialogues that further led to the development of interventions in the community.

## Results

This section outlines the objectives, key activities, stakeholders, and expected outcomes of the interventions implemented in the three communities.

### Alto-Do Cabrito Community

A total of five interventions were developed in Alto do Cabrito (Table 3). The first intervention proposed by the community group involved creating a memorial to document the community’s history. This initiative aimed to challenge the prevailing narratives about the neighbourhood, which often focused on themes of violence and drug trafficking. The memorial project included capturing audiovisual narratives from residents of the neighbourhood. Group members organized themselves to conduct more than 20 interviews with individuals recognized for their significant roles in the community, whether as religious figures, cultural influencers, or social leaders. To facilitate this process, workshops on audiovisual production were conducted, and interview guides were developed. The interviews were recorded, transcribed and edited for inclusion in the audiovisual production. Beyond sharing these videos on a dedicated website, the collaborative efforts have been showcased in a community exhibition. The memorial project has facilitated a re-examination of the neighbourhoods’ narratives, highlighting its identity as a peripheral area while also exploring the connection between present-day health issues and collective memory.

**Table 3:** List of Interventions developed in Alto do Cabrito community.

| Intervention | Aim | Activities | Actors | Frequency | Immediate impact | Long term impact |
| --- | --- | --- | --- | --- | --- | --- |
| Community memorial | Preserve the life story and narratives of residents, their memories, experiences and knowledge about life and health in the territory | Recording interviews and audiovisual production of narratives condensed into 3 documentaries and development of an open repository (i.e., webpage) | Residents (leaders and people of reference in the community) | Continuous | Encouragement and strengthening of bonds and sense of belonging within and between neighbourhoods, contributing to countering stigmatizing and prejudiced hegemonic narratives about the territory. | Strengthen cohesion among community group members and improve intergenerational relationships.<br>Contribute to the construction of health based on the perception of memory as a human experience. |
| Community health fair | Promote integration between communities.<br>Promote awareness about community working groups.<br>Promote health in an expanded way and collaboration between different social, health, artistic and cultural entities | Socialization of preliminary research results, thematic dialogues, provision of health services and carrying out integrative activities (cultural, artistic, poetic displays) | Residents and researchers | Point | Encouraging engagement and building bonds (between residents, communities and research teams); Expanding the communal conception of health | strengthening autonomy and self-management dynamics; strengthening partnerships with other entities |
| Dique Verde Vivo Project | Contribute to the socio-environmental recovery of Dique do Cabrito | Providing awareness about environmental management; Assisting with claims from public authorities; | Community residents and researchers; LIMPURB, SEMOP, and INEMA | Continuous | Improve the aesthetics of the neighbourhood; Reducing exposure to disease vectors; Promoting care for the environment; Valuing art and local | Improve environmental conditions and quality of life for the population living around the Dique, as well as strengthening community cohesion |
|  |  | Implementing environmental and artistic interventions (cleaning the surrounding area and the waters of the Dique) |  |  | artists and strengthening articulation between residents in organizing the event; Integration between communities (Marechal Rondon and Alto do Cabrito)) |  |
| Bota Fora | Promote the removal of unusable waste materials from abandoned spaces | Removal of unusable waste from abandoned spaces with the support of residents and subsequent collection by the public waste agency. | Residents mostly youths, researchers and LIMPURB | Point | Improve the aesthetics of the neighbourhood;<br>Reducing breeding sites for rodents and vectors;<br>Promoting better air quality and improving overall quality of life. | Improved environmental conditions as measured by reduced incidence of vectors, rodents and associated diseases; improved;<br>Improved unity among community groups |
| Artistic paintings (complementary to Bota Fora) | Redevelopment of degraded or abandoned spaces | Mapping of abandoned or degraded spaces and transformation of such places to social hubs using artistic works | Residents mostly youths | Bi-monthly | Improve the aesthetics of the neighbourhood;<br>Promoting the prevention of using renovated spaces as dumping sites | Promote interest in renovating abandoned spaces with artistic work.<br>Improved environmental conditions as measured by reduced incidence of vectors, rodents and associated diseases;<br>Improved unity among community groups |

The second intervention involved organizing a health fair aimed at fostering collaboration among various social, health, artistic, and cultural entities. In partnership with the Federal University of Bahia (UFBA), planning meetings were held to develop dynamic organizational strategies, plan activities, and secure funding. Professionals from municipal and federal sectors, including those in social services, justice, health services, and integrative health practices such as community therapy, massage therapy, and Qi Gong, were consulted to ensure a comprehensive approach. Local artists were also involved, collaborating with residents to develop proposals aligned with the broader health perspective for inclusion in the fair. The health fair venue featured a dissemination tent where the research’s most significant findings were showcased through interactive panels and discussion circles (Table 3).

The third intervention was the “Dique Verde Vivo Project” (literal translation “Living Green Dike project”). It was initiated by the community group, with the aim of restoring and rejuvenating the Dique do Cabrito, an artificial lake that runs through the community’s territory and is linked to the history of socio-cultural development of the neighbourhood. Although the lake is shared between Alto do Cabrito and Marechal Rondon, the idea to restore it emerged because of discussions brought up by the people interviewed for the memorial, as their narratives highlighted the importance of the Dique for the community, but also from the socialization of results, as it highlighted the number of health and socio-environmental issues linked to the Dique. The Living Green Dike project encompasses a range of activities including educational campaigns aimed at raising community awareness for cleaning the lake, tree planting around the lake, a collective action to assemble a ‘Christmas tree’ using recyclable materials, and the creation of a poetic wall. The latter is a cultural activity featuring presentations by around 130 local artists, facilitating the exchange of existing artistic potential within the neighbourhood. To carry out these activities, this project has actively sought partnerships from governmental agencies responsible for (i) refuse collection i.e., LIMPURB- Empresa de Limpeza Urbana de Salvador, (literal translation “Urban Cleaning Company of Salvador”) (ii) SEMOP – Secretaria Municipal de Ordem Publica, (literal translation “Municipal Secretariat of Public Order”) and (iii) INEMA- Instituto Do Meio Ambiente E Recursos Hidricos, (literal translation “Institute of the Environment and Water Resources”) to plan and execute actions. (Table 3)

Fourth, the “Bota Fora” intervention (literal meaning: “to kick out”) aims to improve environmental quality, repel rats and mosquitoes, and enhance aesthetics of neighbourhoods by removing unusable waste materials from abandoned spaces. With the active involvement of residents, particularly youths, researchers, and the public waste agency (LIMPURB), this intervention was designed to clean up abandoned areas and dispose of waste properly. By clearing these spaces, the intervention seeks to reduce breeding sites for rodents and vectors, thereby improving environmental conditions and air quality. The removal of waste also contributes to a better overall quality of life for residents by creating cleaner and more pleasant surroundings. Additionally, this also aims to foster improved unity among community groups as they collaborate towards a common goal of enhancing the neighbourhoods’ cleanliness (Table 3)

Finally, the fifth intervention is complementary to “Bota Fora” and involves redevelopment of degraded or abandoned spaces within the community using artistic paintings. Through collaborative efforts primarily involving youths, these spaces are identified through mapping exercises and transformed into vibrant social hubs adorned with artistic works. By utilizing artistic expression, the initiative aims to improve the aesthetics of the neighbourhood and deter residents from using these spaces as dumping sites. Through the creation of visually appealing areas, residents are encouraged to take pride in and maintain these spaces, fostering a sense of ownership and community pride. Additionally, this initiative contributes to improved environmental conditions by reducing the incidence of vectors, rodents, and associated diseases, thereby enhancing overall public health. Again, the collaborative nature of the project fosters unity among community groups as they work together towards revitalizing their shared spaces and creating a more vibrant and inclusive community environment. (Table 3).

### Marechal Rondon Community

In Marechal Rondon, as the group was primarily made up of young people, the first set of interventions was related to the development of community and university courses and workshops to prepare young people for the job market, considering the greater difficulty in accessing jobs for young people living in peripheries [40]. These activities include courses in English and French languages, computer, robotics, theatre, singing, guitar, dance, college entrance exam preparation classes, entrepreneurship, first-aid and graffiti. One of the results of these interventions was the fact that some young people who participated in these courses were available to share what they learned with other young people in the community. Even after the project ended, these young people continued to teach classes voluntarily in the community and were recognized as teachers. This has brought satisfaction to these young people due to the feedback they have received from their students and due to the difference, they are making in their community. According to the young people, these actions have also helped them create new dreams or have allowed them to dedicate more attention to the dreams they already had. (see Table 4 for details).

**Table 4:**
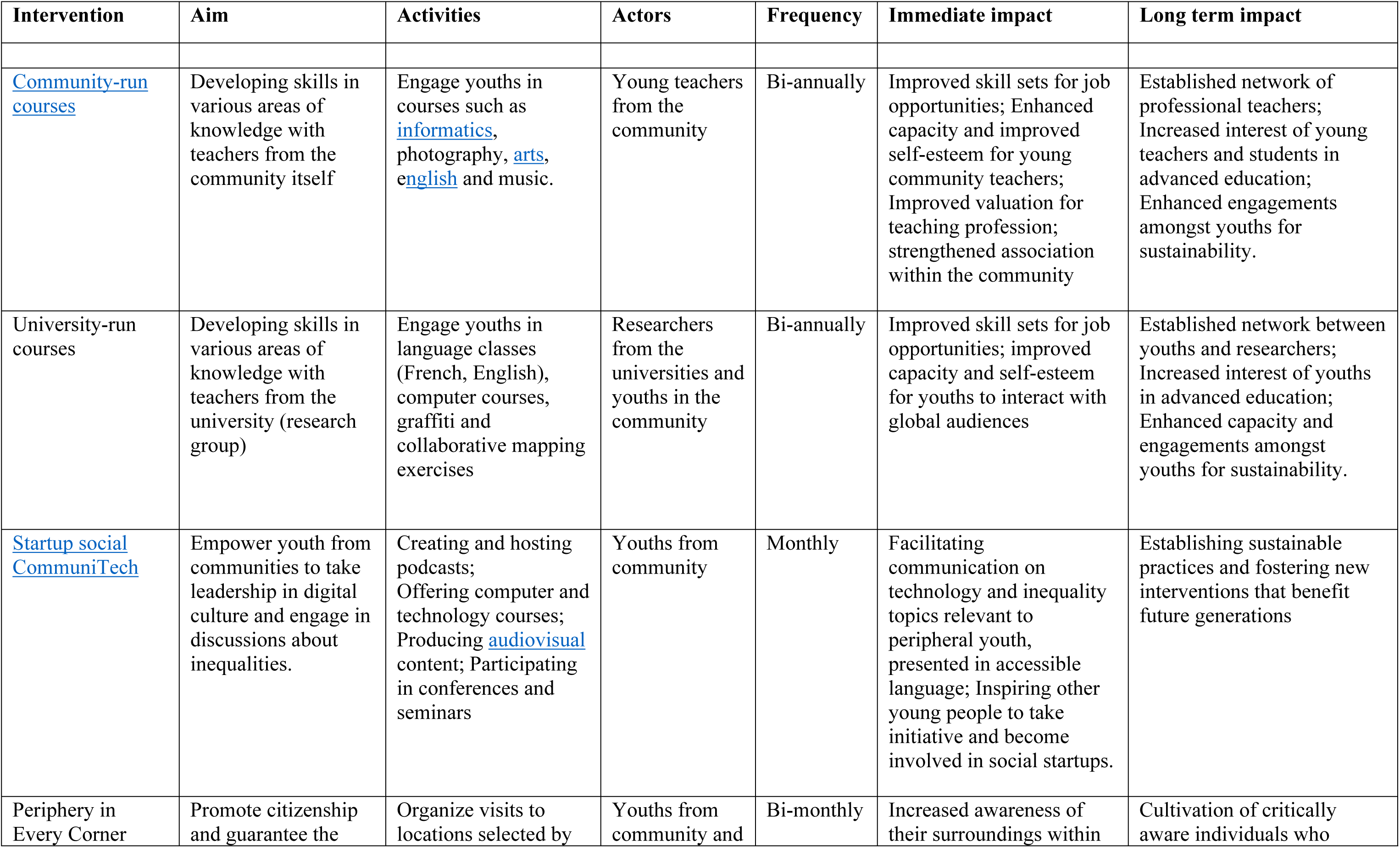
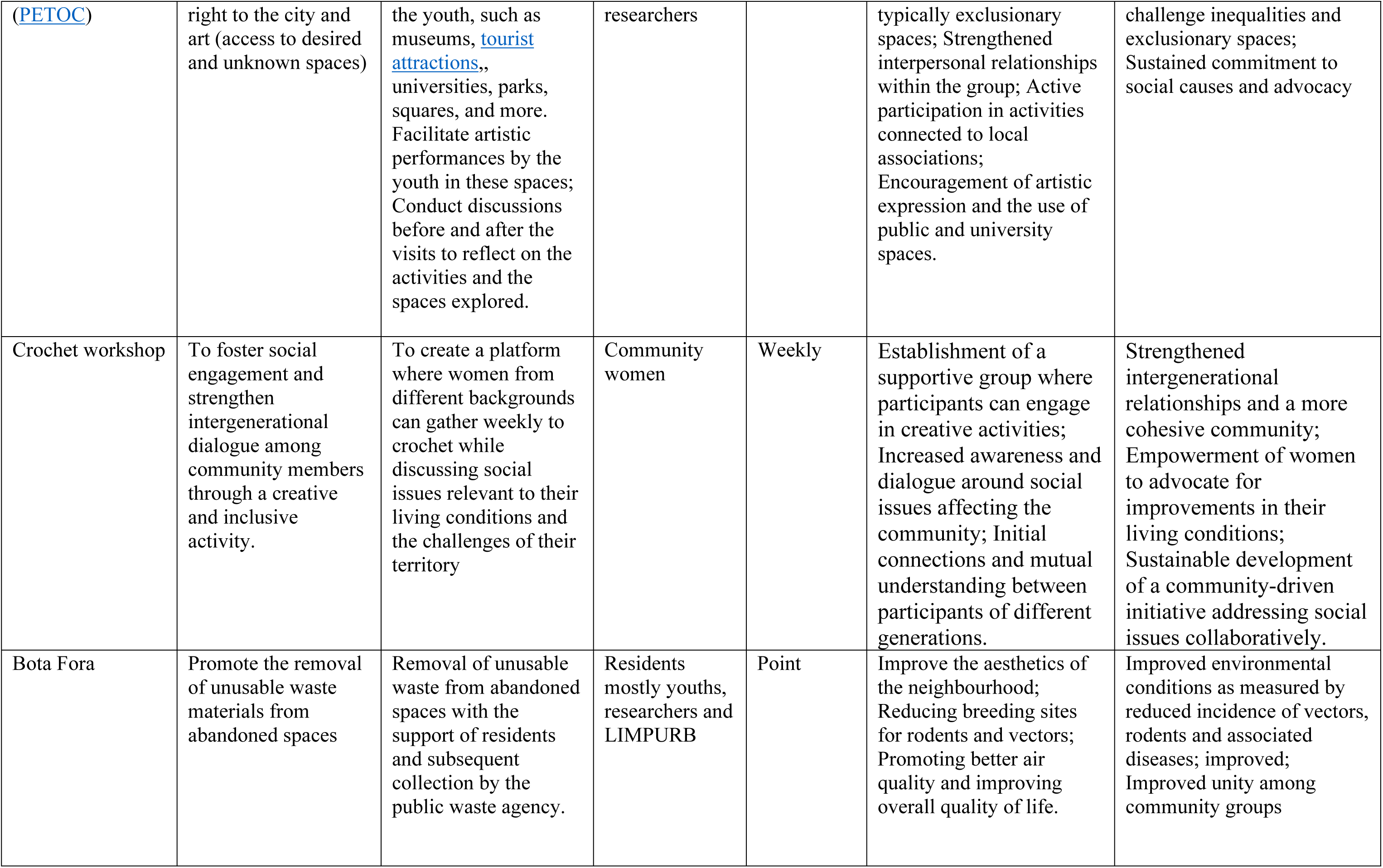
List of Interventions developed in Marechal Rondon community.

The second intervention was “Periferia em Todos os Cantos” (PETOC) project (literal meaning: Periphery in Every Corner), an initiative that enabled young people to occupy spaces in the city that many of them had not yet visited and, in some cases, never considered they could access. The PETOC project promoted artistic performances in various locations in the city, such as the “Universidade Federal da Bahia” (UFBA) and neighborhoods such as Barra and Pelourinho, which are among the most visited places by tourists in Salvador. (see Table 4 for details).

The third intervention, like the memorial intervention in Alto Do Cabrito, was the creation of a podcast to identify and address problems in the community and those identified by the community groups. This initiative seeks to recover memories and expand the possibilities for dialogue in the community on contemporary issues, particularly those related to the environment, inequalities, social empowerment and innovations. This proposal was developed by a group of young people who founded a startup called “Iniciativa Morpheus” (Literal meaning: Morpheus Initiative) with the main objective of continuing to bring improvements to the community after the project is completed through social entrepreneurship practices. With this, these young people seek to give more visibility to the community group, expanding access to the opportunities they had during the project for other residents in the community. (see Table 4 for details).

The fourth intervention was the crochet workshop, which aimed to bring together a different audience from those who already attended the group and to stimulate intergenerational dialogue. Through this initiative, a group of women was formed who began to meet weekly to crochet and discuss social issues related to their living conditions and the territory in which they live. (see Table 4 for details).

Finally, the fifth intervention is the “Bota-fora”, which took place in a similar way to the process developed in Alto do Cabrito, with the two community groups coming together to carry out the activity in partnership. However, In Marechal, there were instances where the researchers interacted extensively with residents and shared information on how to produce homemade repellents to prevent mosquitoes that transmit dengue fever. (see Table 4 for details).

### Pau Da Lima community

A total of four interventions were developed in Pau da Lima. The first, the coverage of sewage, was implemented by the municipal project and not an outcome of our processes. However, the community residents and researchers joined the agency in monitoring and improving the coverage of the intervention. Briefly, the intervention focused on addressing pollution in the Trobogy and Coroado tributaries of the Jaguaribe River by covering sewage channels. The aims are to enhance health conditions and community coexistence dynamics. The other three interventions were designed from the “Freirian tent” during our socialization activity. Events at the tent were coordinated by the community working groups using social cartography methodologies. This allowed the identification of challenges and needs, articulation of community aspirations, and creation of a ‘cartography of dreams’. Residents preferred interventions aimed at strengthening community spaces and bonds beyond the project’s duration. (Table 5).

**Table 5:** List of Interventions developed in Pau da lima community.

| **Intervention** | **Aim** | **Activities** | **Actors** | **Frequency** | **Immediate impact** | **Long term impact** |
| --- | --- | --- | --- | --- | --- | --- |
| *Coverage sewage channels | Cover sewage channels that pollute the Trobogy and Coroado tributaries of the Jaguaribe River | We followed the project, encouraging dialogue between residents and those responsible for the work with the aim of maximizing its scale and benefits | Municipal government agencies. | Point | Improved health conditions and the dynamics of community coexistence. | Improved quality of life |
| Community spaces | Improving the neighbourhoods' infrastructure and expanding access to public services | Construction, and management of parks, playing square, daycare center and family health unit (collectively regarded as eco-social hubs) | Community residents and researchers | Continuous | Improved access to basic services such as parks, playing square, daycare center and family health unit (collectively regarded as eco-social hubs), | Improved quality of life |
| Sports, creative and professional workshops | Promote personal and sociocultural development activities among children and adults within and between communities. | Sporting, recreational and other professional activities targeting children and young people, such as boxing classes, theater and photography workshops | Community residents and researchers | Continuous | Promote strengthening of bonds among children and adults within and between communities; Develop creative and professional capabilities of people in the community; Recognize and discuss the health problems involved in Baixa da Santa Rita | Promote integration among children (for example those with disabilities) and challenge existing social barriers |
| Community garden and kitchen | Promote activities to combat food insecurity and unemployment | Construction of a community garden and kitchen; Establishment of dialogues on food security and income generation | Community residents and researchers | Continuous | Improve food security and relationship with the environment; Improve employment and income conditions, based on practices that encourage solidarity and the construction of citizenship. | Improved quality of life; Improved Food security; Improved income |

\*: interventions were implemented by the municipal government outside the project and were only monitored by the community residents and researchers

First, the need to improve the neighbourhoods’ infrastructure and expand access to public services was identified, which led to the development of spaces that serve both ecological and social functions. This initiative aimed at improving the overall quality of life within the neighbourhood and will involve the construction and management of various amenities, including parks, playgrounds, daycare centres, and family health units. During the project period, the city government covered the sewers and, in addition to residents interacting directly with those responsible for the project to question aspects that they thought could be improved, they also raised questions about paving, spaces for socializing and practicing sports, parks for children, and equipment for exercising. According to those responsible for the project, many of these demands were already foreseen in the project, but these discussions influenced the decision of what was done and where these improvements were implemented. It is notable that this mobilization occurred during the pre-election period, potentially accelerating the completion of the second phase of the project. However, certain community needs, such as the establishment of daycare centers and a family health unit, remain unmet. Nevertheless, residents continue to seek ways to improve access to health services and to have daycare centers for children in the community itself. (Table 5).

The second group of interventions was the development of sporting, recreational and other professional activities targeting children, young people and adults. This process included activities such as boxing, theatre, photography, crafts and makeup workshops to promote integration among the community’s residents and to challenge existing social barriers, as well as courses aimed at expanding the possibilities for young people to enter the job market. The theatre workshops were structured to encompass training modules covering dance, theatrical interpretation, artistic makeup, and music, with an emphasis on fostering ethics, critical thinking, and aesthetics. Similarly, the boxing workshops were crafted to integrate boxing training within a healthcare context, complemented by discussion circles that intersect with other project activities. Additionally, the photography component aimed to involve participants in activities such as sharing technical and aesthetic aspects of photography, enabling interested individuals to contribute to participatory research endeavours. The makeup course (coordinated by a resident), in addition to sharing beauty techniques, was created to be a space to discuss self-care and socially imposed beauty standards. Finally, the crafts course (coordinated by a resident) was attended by adults and children, sharing different techniques, mainly using recyclable materials to produce art. (Table 5).

The third intervention is a community garden and kitchen, which was designed for young people and adults, emphasizing alternatives for income generation while addressing food insecurity issues and improving nutrition for residents facing financial constraints. It involves the construction of the community garden and kitchen itself, but also the establishment of regular dialogues on food security and income generation in the community (Table 5).

## Discussion

In this article, we have demonstrated how our interdisciplinary program could leverage social capital and inter-sectoral partnerships to enhance the capacity of marginalized urban communities to address health and environmental challenges through locally designed interventions that are sustainable and cost-effective [41]. In the context of participatory research, particularly those addressing health challenges in informal urban settlements, our program introduces several innovative dimensions that contribute to the field. Foremost, working simultaneously in three communities allowed integration of diverse knowledge, perspectives, and priorities, with a particular focus on engaging women and youth. Secondly, the communities identified priorities and targets based on their specific cultural, social, and environmental perspectives. Thirdly, the unique interventions implemented by each community to tackle similar problems demonstrated that optimal solutions are context-dependent rather than universally ‘best’. Furthermore, from the outset, our team prepared to evaluate changes in incidence of specific infectious diseases and environmental indicators as metrics to accompany the interventions. Interestingly, the communities primarily chose multi-purpose interventions targeting general wellbeing—choices that differed from our expectations based solely on previously collected eco-epidemiological data [10]. This unexpected preference highlighted the importance of addressing broader social and collective determinants of health and will allow us to test whether the benefits of these holistic interventions extend to more specific outcomes, such as reductions in single diseases or improvements in environmental indicators [42,43]. Another innovative aspect was implementing a feedback loop connecting the *activities* organised by our team and grounded in Popular Health Education [44] with the *interventions* initiated by the community. This dynamic facilitated continuous dialogue and mutual adaptation, enabling us to refine our activities and research instruments both within and across disciplines.

Below, we describe these dimensions in more detail and provide examples from the three communities where the program was implemented. We also describe how community participation led to setting joint agendas and plans with governmental agencies, in addition to secondary and medium- and long-term benefits for the community. Finally, we identify some of the challenges that our program faced and the lessons that can be learned for adapting such a program to other contexts.

### Unique solutions to common challenges

Our program embraced the diverse knowledge and perspectives both within and among communities, drawing from differences in demographic, socioeconomic, and cultural characteristics of existing organizations, newly formed community groups, and participating residents. While the three communities face similar health and quality of life challenges (Table 1), their distinct profiles have shaped local priorities, resources, and methods of engagement, leading to a diversity of solutions tailored to their specific needs [28]. This aligns with earlier studies emphasizing that context-specific and inclusive interventions are more effective than one-size-fits-all models [45,46]. By remaining attentive to the unique journey of each community, including their distinct histories, starting points, and processes, we aim to understand, upon evaluating the interventions in the next phase of the program, how the local context can influence priorities, interventions, and their outcomes. This understanding will highlight key similarities and differences among the communities, which will aid adapting the program to other contexts nationally and internationally [48].

In particular, the active participation of women and youth introduce diverse perspectives and fosters innovative approaches that are more attuned to the broader needs of the community [49]. Despite their important role, these groups are frequently underrepresented in participatory initiatives, leading to gaps in addressing their specific needs and perspectives [50]. Involving women in community health initiatives has been shown in some contexts to improve health outcomes, as they are often primary caregivers and influencers in their communities [51,52]. Similarly, youth engagement fosters innovation and ensures that interventions are aligned with the needs of younger generations [53]. Their involvement often results in interventions that prioritize social determinants of health and strengthen community ties and capacity, bringing forward concerns and ideas that might otherwise be overlooked. These include gender-specific health needs, youth engagement strategies, child welfare, cultural preservation, and innovative approaches to community development [49]. In the community of Pau da Lima, most participants were women. They highlighted social vulnerabilities related to gender, class, and ethnicity, such as food insecurity, violence, and mental health. These themes were closely linked to the interventions proposed afterwards, which included a community garden and kitchen, sports and artistic activities, and workshops (e.g., crafts and makeup).

Even when similar interventions were adopted in the two neighboring communities of Marechal Rondon and Alto do Cabrito, there were differences in how and when these interventions were framed. For instance, Marechal Rondon chose to document their history through a youth- and technology-driven podcast initiative, drawing inspiration from Alto do Cabrito, which had initially preserved its history through memorials. Similarly, the revitalization of the lake and surrounding area was a joint proposal by the extension team to both communities, following complaints about garbage accumulation. While the implementation models varied, particularly in the mobilization of actors, both communities adopted the same partnership dynamic: collecting materials as feasible, placing them in designated locations, and having LIMPURB handle subsequent removal.

Thus, our program moves beyond the single-community and single-approach studies prevalent in much of the existing CBPR literature [54]. Moreover, by examining the distinct starting points, processes, and outcomes of each community, our program can aim to discern, following co-evaluation, which common factors contribute to successful interventions. This, in turn, can inform future projects and policies, and will be crucial for scaling up interventions while ensuring they are culturally sensitive and contextually appropriate [54].

### Evaluating broad interventions with a wide range of metrics

Rather than imposing predefined intervention targets—which often limits participation and local input into not only *what* solutions are prioritised, but also *how* they are implemented [48,55], our program facilitated interventions that emerged organically from the concerns of the involved communities. The challenges identified spanned environmental, individual, and community levels—including infectious diseases, physical and mental wellbeing, food security, and violence—as well as intangible elements like memory, identity, and sense of community. Interestingly, the interventions chosen in all three communities overwhelmingly targeted general wellbeing—choices that differed from what we might have selected as researchers relying solely on eco-epidemiological data.

From the outset, our program was prepared to continuously update existing evaluation metrics and introduce new ones, since we anticipated that the diverse community-driven processes would lead to a wide array of initiatives addressing both specific diseases and general wellbeing. We adopted a systems approach to evaluating these complex interventions to acknowledge that addressing broad, overarching wellbeing goals can have ripple effects on specific outcomes such as disease prevalence and environmental health [56]. This holistic perspective therefore allows us to assess whether broad interventions carry over to specific health outcomes, collaboratively defined at individual, community, animal, and environmental levels, as well as to stakeholder participation and satisfaction.

This approach moves beyond the limitations of traditional projects that use CBPR only as an instrument and therefore focus on narrow, predefined variables [57]. Instead, our program will define intervention success not only through individual-level outcomes (e.g., disease incidence) and at community, animal, and environmental levels, but also, more importantly, by fully appreciating community health and empowerment [54]. Through evaluating interventions within the complex systems they operate in, we will attempt to capture the interconnectedness of various health determinants [58]. This includes revealing whether interventions targeting individual and community wellbeing, shaped by their choices, can yield specific health benefits, and challenges the assumption that the most effective interventions are those defined solely by a narrow set of predefined outcomes. For example, in Alto do Cabrito, the decision to create a memorial to reclaim the identity and history of the community became a catalyst for further interventions, including reclaiming a dumpsite as a football field in the “Bota Fora” intervention which was also implemented in Marechal Rondon. This transformation is expected not only to enhance physical and mental health but also to reduce inadequate waste disposal, potentially decreasing rat presence and diminishing the transmission of leptospirosis—a key infectious disease in the community [10]. In Pau da Lima, sports and creative workshops for children extended beyond traditional health concerns by addressing social integration and personal development. These projects spurred increased participation by youth and their mothers in broader health initiatives, leading to the creation of a community garden and kitchen that addressed food insecurity while providing environmental and economic benefits, potentially reducing disease risks (by removing resources for rats and mosquito breeding sites), and fostering community resilience.

### Interdisciplinarity of focus

Throughout the program, our research teams needed to be adapted to these multi-target and interlinked interventions by stepping out of narrow disciplinary discourses and processes and sharing experiences to formulate new research tools. Regular meetings and workshops, involving community representatives, were key to synthesising diverse disciplinary and interdisciplinary perspectives, ensuring that the insights from one team informed and enriched the methodologies and approaches of the others. As one example, the “Dream Mapping” integrated social science activities with remote-sensing and mapping efforts. As another, if residents indicated a hotspot of disease transmission, the epidemiology and ecology teams would adjust their sampling to focus intensively on that area, while social scientists conducted in-depth interviews with the residents to explore factors contributing to exposure risk and barriers to avoiding contact with sources of contamination.

The teams and the program also benefitted from the interplay between *activities* and *interventions*. A*ctivities* aimed to engage and mobilize the community and were jointly undertaken by the research team and members of the communities themselves, acting as mobilisers and co-researchers. This feedback loop between *activities* and *interventions* culminated in interventions that were more creative and multi-targeted than what would have likely been proposed by a more conventional approach, driven mainly by formally trained researchers, [59–61], or with an approach where both researchers and community cross-fertilized ideas but were limited within a fixed scope of intervention targets and outcomes [62,63]. Thus, the flexible approach by our team and the interplay between *activities* and *interventions* resulted in more interdisciplinarity in our program and incentivised our program the expansion of intervention targets and outcomes to include multiple health- and wellbeing-related issues.

### Program short and long-term benefits

In the interim, our approach has not only succeeded in designing and implementing interventions to address health concerns as perceived by the community residents, it has also been instrumental in promoting within underrepresented communities the possibility of participation in local planning and governance, which is uncommon in peripheral communities [64], and also the possibility of setting joint priorities and agendas with governmental agencies, with three hearings made between 2022-2024 on “Bota Fora”, and “Living Green Dyke” interventions that required continuous collaboration with three agencies (LIMPURB, SEMOP, and INEMA). Furthermore, it has fostered community unity, the establishment of common ground, and the emergence of leaders from historically marginalized groups. These leaders play a crucial role in both the implementation and sustained maintenance of interventions.

Nevertheless, the need for robust evaluation of our program impacts, assessed through both implementation outcomes and the effects of interventions, has become increasingly critical [65]. Implementation outcomes specifically measure the success of the implementation process itself, distinct from the anticipated effects or impact of the intervention after a designated period. This distinction is vital for driving sustainable changes at local scales. When interventions fail to meet expectations, it is essential to discern whether the failure was due to the intervention being ineffective (intervention failure) or the improper execution of an otherwise effective intervention (implementation failure). Evaluating implementation effectiveness involves assessing key factors such as acceptability, appropriateness, feasibility, uptake (adoption), reach (penetration), fidelity, cost, and sustainability. These metrics provide valuable insights to refine the implementation process and maximize its impact. Importantly, interventions in informal settlements hold significant public value, even when their effects are not immediately measurable using conventional epidemiological or ecological metrics—challenging the perspective we held during the project’s conceptualization. Hence, it is imperative to prioritize the evaluation of both immediate (short-term) outcomes and the long-term impacts of these interventions on broader health determinants.

### Challenges

Our CBPR process faced significant resource and timing challenges like those encountered in previous studies [54, 59, 60, 61]. More specifically, communities lacked existing groups and representative associations, suffered from fragile social cohesion, and faced violence. In Pau Da Lima, the absence of community spaces hindered socialisation, prompting the creation of “Freirean Tent” activities, which eventually led to the formation of the United Community Group, a name chosen by participants themselves to emphasize the importance of unity for achieving improvements, rights, and health outcomes. Physical space limitations impacted interventions like boxing and theatre workshops, which limited children participation. Adverse weather and insecurity, including police operations, further disrupted activities, highlighting the community’s struggle with invisibility and the incomplete implementation of established rights. For the research team, building trust within teams and communities, embracing the dynamic nature of CBPR, and effectively integrating interdisciplinary approaches provided challenges but also valuable opportunities for growth and collaboration. Conducting participatory research during the pandemic also added complexity, limiting interactions to virtual meetings over a long period of the project. Defining the scope of participation and ensuring effective co-construction of knowledge was also difficult, and the team had to expand their health concepts to align with local realities and re-assess protocols and “timelines” multiple times. Diverse methods and engagement modes made controlling the research trajectory challenging but encouraged dialogue among stakeholders, avoiding the imposition of a singular “truth.”

## Conclusion

This article outlines a Community-Based Participatory Research (CBPR) approach, combined with Popular Health Education principles, to address health challenges in neglected urban communities in Salvador, Brazil. The approach stands out by engaging diverse community members, considering their unique perspectives based on gender, age, and social status, and addressing multiple health-related issues beyond infectious diseases, such as food security, violence, memory, and identity. By integrating local knowledge with flexible, interdisciplinary methods, the project led to community-driven interventions that were tailored to the specific needs and aspirations of the residents, fostering community unity and inclusivity, particularly among women, children, and youth. Despite challenges such as resource limitations, violence, and the COVID-19 pandemic, the project succeeded in promoting participation in local planning, establishing common ground, and empowering marginalized groups to take leadership roles. The need for further evaluation of both the short-term and long-term impacts of these interventions is emphasized to ensure their sustainability and effectiveness.

## Author Contributions

Conceptualization: HK, MB, FC, LYAA, HM; Methodology: HK, MB, FC, LYAA, HM, LAF, JHAV, MAA, HA, IS, APM, TL, SC,RB, EC, ECNS, EL, ER, TB,TA, NNJ, JC, IC, RL, VD, MR, AIK; Software: HM, HA, NNJ, JC, JHAV, IC, RL, VD, MAA, HK; Validation: MB, FC, LYAA, HK, MR, AIK; Formal Analysis: HK, MB, FC, LYAA, HM, LAF, JHAV, MAA, HA, IS, APM, TL, SC, RB, EC, ECNS, EL, ER, TB,TA, NNJ, JC, IC, RL, VD; Investigation: HK, MB, FC, LYAA, HM, LAF, JHAV, MAA, HA, IS, APM, TL, SC,RB, EC, ECNS, EL, ER, TB,TA, NNJ, JC, IC, RL, VD, MR, AIK; Data curation: HM, HA, NNJ, JC, JHAV, IC, RL, VD, MAA, HK Writing—original draft preparation: HM, HK, MB, JHAV, HA; Writing—review and editing: HK, MB, FC, LYAA, HM, LAF, JHAV, MAA, HA, IS, APM, TL, SC,RB, EC, ECNS, EL, ER, TB,TA, NNJ, JC, IC, RL, VD, MR, AIK; Visualization: HA, HM, NNJ, RL, HK; Supervision: MB, FC, LYAA, HK, MR, AIK; Project administration: HM, HA, NNJ, JC, JHAV, IC, RL, VD, MAA, HK; Funding acquisition: MB, HK, FC; All authors have read and agreed to the published version of the manuscript.

## Institutional Review Board Statement

This study involves human participants and was approved by the Research Ethics Committee of the Institute of Collective Health/Federal University of Bahia (CEP/ISC/UFBA) and a National Research Ethics Committee (CONEP) linked to Brazilian Ministry of Health (approval number 35405320.0.1001.5030). All participants were informed about the study procedures and agreed to participate by signing a written informed consent form (for adults above age 17) or an assent form (for minors below age 18). Participants gave informed consent to participate in the study before taking part.

## Informed Consent Statement

Informed consent was obtained from all subjects involved in the study.

## Data Availability Statement

All relevant data used in this manuscript are included in the form of tables or text descriptions. Additional datasets generated through this project can be provided upon reasonable request from the Data Manager at.

## Acknowledgments

The authors would like to acknowledge Emilia Machado, the study participants, communities, and all academia who provided support in the design and implementation of this project.

